# DNA methylation age acceleration is associated with incident cognitive impairment in the Health and Retirement Study

**DOI:** 10.1101/2024.09.19.24314012

**Authors:** Freida Blostein, Kelly M. Bakulski, Mingzhou Fu, Herong Wang, Matthew Zawistowski, Erin B. Ware

**Affiliations:** Department of Epidemiology, School of Public Health, University of Michigan; Department of Medical Informatics, University of California, Los Angeles; Department of Biostatistics, School of Public Health, University of Michigan; Survey Research Center, Institute for Social Research, University of Michigan

**Keywords:** DNA methylation age, epigenetics, cognitive impairment, dementia, cohort

## Abstract

**Background:** DNA methylation clocks have emerged as promising biomarkers for cognitive impairment and dementia. Longitudinal studies exploring the link between DNA methylation clocks and cognitive decline have been constrained by limited sample sizes and a lack of diversity.

**Objective:** Our study aimed to investigate the longitudinal associations between DNA methylation clocks and incident cognitive impairment using a larger sample size encompassing a US nationally representative sample from the Health and Retirement Study.

**Methods:** We measured DNA methylation age acceleration in 2016 by comparing the residuals of DNA methylation clocks, including GrimAge, against chronological age. Cognitive decline was determined by the change in Langa-Weir cognition status from 2016 to 2018. Using multivariable logistic regression, we evaluated the link between DNA methylation age acceleration and cognitive decline, adjusting for cell-type proportions, demographic, and health factors. We also conducted an inverse probability weighting analysis to address potential selection bias from varying loss-to-follow-up rates.

**Results:** The analytic sample (N=2,713) at baseline had an average of 68 years old, and during the two years of follow-up, 12% experienced cognitive decline. Participants who experienced cognitive decline during follow-up had higher baseline GrimAge (mean = 1.2 years) acceleration compared to those who maintained normal cognitive function (mean = -0.8 years, p < 0.001). A one-year increase in GrimAge acceleration was associated with 1.05 times higher adjusted and survey-weighted odds of cognitive decline during follow-up (95% CI: 1.01-1.10). This association was consistent after accounting for loss-to-follow-up (OR = 1.07, 95% CI: 1.04-1.11).

**Conclusion:** Our study offers insights into DNA methylation age acceleration associated with cognitive decline, suggesting avenues for improved prevention, diagnosis, and treatment.

## Introduction

The aging population structure in the United States (US) is leading to an increasing public health challenge, especially with age-related diseases like dementia. In 2020, it was estimated that around 6.07 million individuals in the US had clinical Alzheimer’s disease, and another 12.23 million exhibited mild cognitive impairments [1]. By 2060, these figures are projected to rise to 13.85 million and 21.55 million, respectively [1]. Both Alzheimer’s disease and cognitive impairments severely restrict an individual’s quality of life and necessitate substantial caregiving. Despite extensive research, there are only a few effective treatments available, and those that do exist seem to work best when administered early in the progression of the disease [2]. Therefore, pinpointing those at high risk for developing dementia or cognitive impairments is crucial for suggesting preventive measures and initiating timely interventions.

A promising biomarker for cognitive impairment and dementia is epigenetic aging. Rather than altering the DNA sequence itself, epigenetic factors like histone modifications and DNA methylation modify the regulation and expression of DNA. Among these, DNA methylation patterns have been particularly adept at forecasting chronological age and mortality risks [3]. By integrating DNA methylation levels from multiple genomic sites, DNA methylation clocks attempt to provide a more accurate depiction of biological aging [4]. Residualizing DNA methylation clocks against chronological age creates a measure of whether an individual’s DNA methylation age is older (accelerated aging) or younger (not accelerated) than would be expected [5,6]. While the initial versions of DNA methylation clocks aimed to predict chronological age [7,8], the second generation, often termed ‘phenotypic clocks,’ are designed specifically to forecast age-related health decline [7–9]. Consequently, these updated clocks might serve as more reliable biomarkers for cognitive deterioration and dementia.

Prior research has explored the associations between DNA methylation clocks and outcomes related to dementia and cognition, yet the findings have been inconclusive [10]. A recent systematic review identified 10 studies examining associations between DNA methylation clocks and dementia, including Alzheimer’s disease, cognitive impairment, and frontotemporal dementia [10]. Among these, only four had a longitudinal design, with just one reporting a significant finding [11–14]. Moreover, the sample sizes of these longitudinal studies were smaller, ranging from 52 to 578 participants [10]. In contrast, larger cross-sectional studies (ranging from 29 to 4535 participants) investigating DNA methylation age and cognitive metrics have yielded stronger associations, particularly with second-generation clocks like GrimAge [10]. However, for DNA methylation age acceleration to function as a viable biomarker, differences must be preceding clinical cognitive decline.

To bridge these knowledge gaps, we examined the relationships between DNA methylation clocks and the onset of cognitive decline. This analysis involved 2,713 participants from the Health and Retirement Study (HRS), a nationally representative longitudinal cohort study focused on older adults in the US.

## Materials & Methods

### Study population and design

This study is based on data collected from the 2016 and 2018 waves of the HRS. The HRS, funded by the National Institute of Aging and the Social Security Administration, is a longitudinal panel study of older adults (>50 years of age) in the US, with sample replenishment every six years with younger cohorts [15]. HRS collects demographic, economic, and health metrics every two years, with participants alternating between face-to-face interviews and telephone surveys, such that each participant completes a face-to-face interview every 4-years. In 2016, all community-dwelling HRS participants who completed an interview and did not respond via proxy were asked to consent to a venous blood draw. A total of 9,934 participants consented. From these, a subsample (N = 4,104) was selected for DNA methylation assays. We considered DNA methylation measures in 2016 as the study baseline and we followed-up cognitive status into the 2018 wave (most recent available). All participants in the HRS provide informed consent. This secondary analysis was approved by the University of Michigan Institutional Review Board (HUM00128220).

### DNA methylation age measures

The primary exposure variable for this analysis was DNA methylation age acceleration. DNA methylation was measured using the Infinium Methylation EPIC BeadChip at the University of Minnesota. To minimize batch confounding, samples were randomized across plates by demographic variables. To assess technical variability, 40 pairs of blinded technical replicate samples were included; technical replicates showed high correlation (>0.97) across all DNA methylation sites. DNA methylation data was preprocessed by HRS staff using the *minfi* package in R [16]. Briefly, DNA methylation probes with a detection P-value below 0.01 were excluded from the final data set. Samples which failed preprocessing (detection P-value 5% threshold) or had a mismatch between DNA-methylation sex and reported sex were excluded, leaving 4,018 samples with quality-controlled DNA methylation data. Prior to estimation of DNA methylation clocks, the HRS imputed missing DNA methylation probe values using the mean value of the given probe across all samples.

Thirteen epigenetic clocks were constructed by the HRS. We focused on five widely recognized epigenetic clocks based on *a priori* hypotheses. Our primary interest was in three phenotypic clocks: GrimAge [6], DunedinPoAm (MPOA) [17], and PhenoAge [18]. As a secondary analysis, we also included two chronological clocks: Horvath [19] and Hannum [20]. Four of the clocks provided DNA methylation age estimates in years. MPOA was provided as a rate, and we transformed it to years by dividing by the participant age to be consistent with the remaining clocks [17].

To account for chronologic age and compute DNA methylation age acceleration, we regressed each clock against the chronological age at the time of measurement, and we extracted the residuals [5,6]. We visualized patterns in participant measures in the clocks, chronologic age, and DNA methylation age acceleration (residuals) using scatter plots and computed Pearson correlation coefficients (**Supplemental Figure 1**). We examined the pairwise correlation between five epigenetic clocks using a Pearson correlation plot (**Supplemental Figure 2**). In descriptive analyses, we dichotomized accelerated DNA methylation ages (residuals greater than 0) and non-accelerated DNA methylation ages (residuals 0 or less).

### Cognitive measures

Cognition was measured at baseline in 2016 and at follow-up in 2018 using the Langa-Weir three-level cognition status (normal cognition, cognitively unimpaired non-dementia, and dementia) [21]. Each wave, the HRS team provides imputed values for missing cognitive tasks, which are then included in an overall 27-point score [22]. The Langa-Weir cognition status is calculated differently for self-respondents and proxy respondents. No proxy respondents were included in the baseline wave, though 23 respondents transitioned to proxy status due to cognition concerns in 2018. Of these 23 respondents, nine were categorized as having dementia, and seven each were classified as cognitive impairment, non-dementia and normal cognition. in For self-respondents, a 27-point cumulative cognition score based on four cognitive tasks administered to participants is used for classification: immediate and delayed 10-noun free recall task (0-20 points), serial sevens subtraction task (0-5 points), and backward counting task (0-2 points). The 27-point summary score is then categorized into normal cognition (12-27 points), cognitive impairment non-dementia (7-11 points) and dementia (0-6 points). While no participants from the 2016 baseline venous blood draw sample were proxy respondents, 23 respondents transitioned to proxy status in the 2018 wave. For those proxy respondents, Langa-Weir cognition status is determined using an 11-point scale, constructed using a proxy’s assessment of the subject’s memory (ranging from excellent to poor), limitations in instrumental activities of daily living, and an interviewer’s assessment of the subject’s cognitive impairment. Higher scores indicated worse cognition. As with the score for self-respondents, missing data is addressed through imputation. The aggregate 11-point score is then classified into normal cognition (0-2 points), cognitive impairment non-dementia (3-5 points), and dementia (6-11 points) [21]. The cognitive status measure that has been clinically validated with a 74% sensitivity [21].

We visualized transitions between cognitive states between baseline and follow-up using an alluvial plot. For analyses of cognitive decline, we excluded participants with dementia at baseline, since they could not undergo any further cognitive decline, as well as those with a stable, non-normal cognitive conditions. To examine cognitive decline between baseline and follow-up, we categorized participants who underwent any decline in Langa-Weir cognitive status between 2016 and 2018 -- specifically, transitions from normal to impairment, normal to dementia, or impairment to dementia. This category was contrasted with participants who consistently maintained a cognitively normal status (remained cognitively normal from 2016 to 2018) or improved cognition.

### Covariate measures

We sought to identify potential confounders and precision variables. Considering that each cell type possesses its own distinct DNA methylation profile, which can be associated with health outcomes, it’s crucial to adjust for cell type distributions [23]. To this end, we used the proportions of granulocytes, lymphocytes, and monocytes in venous blood samples from HRS participants measured using complete blood counts.

We accounted for various demographic variables, including chronological age in 2016 (years), self-reported race (Black/African-American, White, or Other), self-reported Hispanic ethnicity (Hispanic or not Hispanic), sex (male, female), years of education, and marital status in 2016 (single or married/partnered).

We also integrated several health-related factors: cigarette smoking status in 2016 (never or ever smoker), any alcohol consumption in 2016 (any or none), physical activity in 2016 (indicator variable for any self-reported light, moderate or vigorous physical activity more than once per week, with its structure echoing the World Health Organization’s guidelines for seniors [24]), body mass index (BMI) calculated from self-reported height (feet and inches) and weight (pounds), and an indicator variable reflecting whether a participant had been diagnosed by a physician with multiple comorbid conditions (including high blood pressure, diabetes, cancer, lung disease, heart disease, stroke, psychiatric problems, and stroke). Given that many in this aging cohort reported at least one health condition, this indicator was defined as more than one versus one or no comorbid conditions.

The apolipoprotein E (*APOE*) gene is a recognized genetic risk factor for late onset Alzheimer’s disease. Specifically, individuals carrying the *APOE* ϵ4 allele face a heightened risk for dementia compared to those with the *APOE* ϵ3 and *APOE* ϵ2 alleles [25]. Between 2006 and 2012, participants in the HRS underwent face-to-face interviews, during which they provided saliva samples for genotyping. These samples were analyzed using the Illumina Human Omni2.5 microarray platform and later imputed with the 1000 Genomes Project reference panel [26].

Given the rarity of individuals possessing two copies of *APOE* ϵ4, participants were categorized as having any versus no copies of *APOE* ϵ4 using the phased genetic data. However, a proportion of participants lacked genetic data, thus we included *APOE* ϵ4 status as a variable in our sensitivity analysis.

### Sampling weights

We applied the HRS survey weights specifically designed for use with DNA methylation data [27]. We used these sample weights for both descriptive statistics and multivariable models to ensure our findings are nationally representative.

### Sample inclusion and exclusion

Only participants with quality-controlled DNA methylation data from the 2016 venous blood sample were eligible for inclusion in this analysis, and analyses were therefore necessarily restricted to individuals who 1) were community-dwelling and 2) did not respond via proxy in 2016. Participants could, however, be proxy-respondents in 2018. We performed a complete case analysis, where we excluded participants lacking cell type data, demographic information, or health factors variables. Those classified with dementia in 2016 were excluded since they couldn’t experience further cognitive decline. Anyone without cognition data from the 2018 HRS wave was also excluded, as we couldn’t evaluate their cognitive decline. Additionally, participants with non-normal cognition that remained stable or improved between 2016 and 2018 were also excluded. We visualized participant exclusion and inclusion using a flow chart.

### Statistical analysis

All analyses were conducted in R statistical software (version 4.2.2). We described the distributions of continuous variables using mean and standard deviation and of categorical variables using count and frequency. We compared the distributions of variables in the included and excluded samples. For bivariate analysis among the included sample, we took survey weight into account and compared the distributions of variables by binary cognitive decline status using

Wilcoxon rank-sum test for complex survey samples to assess mean differences in continuous variables and Pearson’s Chi-squared test with Rao & Scott’s second-order correction for categorical variables. In sensitivity analyses, we examined subtypes of cognitive decline (normal to cognitive impairment non-dementia, normal to dementia, and cognitive impairment non-dementia to dementia). Raincloud plots were used to visualize the distribution and provide summary statistics of the DNA methylation age acceleration by cognitive decline.

To assess the relationship between DNA methylation age acceleration and cognitive decline, we used multivariable logistic regression, accounting for potential confounders and precision variables. Based on *a priori* hypotheses, our primary multivariable models focused on continuous GrimAge acceleration and the remaining clocks were used in sensitivity analyses.

Our primary analysis encompassed three distinct regression analyses: 1) base model: controlled for cell-type proportions; 2) demographic model: adjusted for demographic variables previously mentioned, in addition to cell-type proportions; and 3) health factors model: adjusted for the health-related factors specified earlier, in addition to all variables included within the demographic model. We also assessed if chronological age modified the association between DNA methylation age acceleration and cognitive decline by adding product term between chronological age and DNA methylation age acceleration in the demographic and health factors models. To better represent those participants who had complete data in 2016, we applied survey weight to these models. We reported odds ratios (OR), 95% confidence intervals (CI), and p-values for the variables of interest.

### Sensitivity analyses

In sensitivity analyses, we considered a binary GrimAge acceleration predictor. In addition, to determine whether the association between DNA methylation age acceleration and cognition decline was specific to the GrimAge estimator, we examined four alternate DNA methylation age estimators (continuous and binary) including Levine, MPOA, Horvath, and Hannum. To address potential confounding by the *APOE* genotype, we additionally controlled for any *APOE* ϵ4 alleles in a sensitivity analysis. All these models were survey sampling weighted.

To address the possible selection bias caused by differing loss-to-follow-up rates between 2016 and 2018, we conducted an inverse probability weighting analysis. First, we compared the distribution of variables among participants who had baseline DNA methylation and cognition measures but did not have cognition measures at follow-up (loss-to follow-up sample) to the analytic sample (that had cognition measures at follow-up). Next, we calculated the inverse probability for both treatment (referring to DNA methylation age acceleration) and censoring (specifically for participants without cognitive data in 2018). For our sensitivity analysis, we multiplied the inverse probabilities of treatment and censoring weights by the given sampling weights. These adjusted weights were then applied to our multivariable models.

To assess if GrimAge acceleration provides increased predictive accuracy beyond standard demographic variables in predicting future cognitive decline, we performed an area under the receiver operator curve analysis (AUROC) using the *pROC* package (version 1.18.0) [28]. We considered our base model to be a multivariable logistic regression model of cognitive decline status with demographic and cell type variables as predictors. We added GrimAge acceleration to this model and tested for an association using a likelihood ratio test. We visualized these results using a receiver operator curve, calculated C-statistics for each model, and used a DeLong test to compare performance.

## Results

### Description of the analytic sample

Of the initial 4,104 participants tested with DNA methylation assays, 4,018 met the HRS quality control standards. After accounting for those with incomplete covariate data, the sample was reduced to 3,713 participants. From this group, 439 were excluded because of stable non-normal cognition, and another 417 lacked cognition data for 2018. Ultimately, 2,713 participants were included in the final analysis (**Figure 1**). During follow up, 333 participants experienced some form of cognitive decline (**Supplemental Figure 3**).

**Figure 1.**
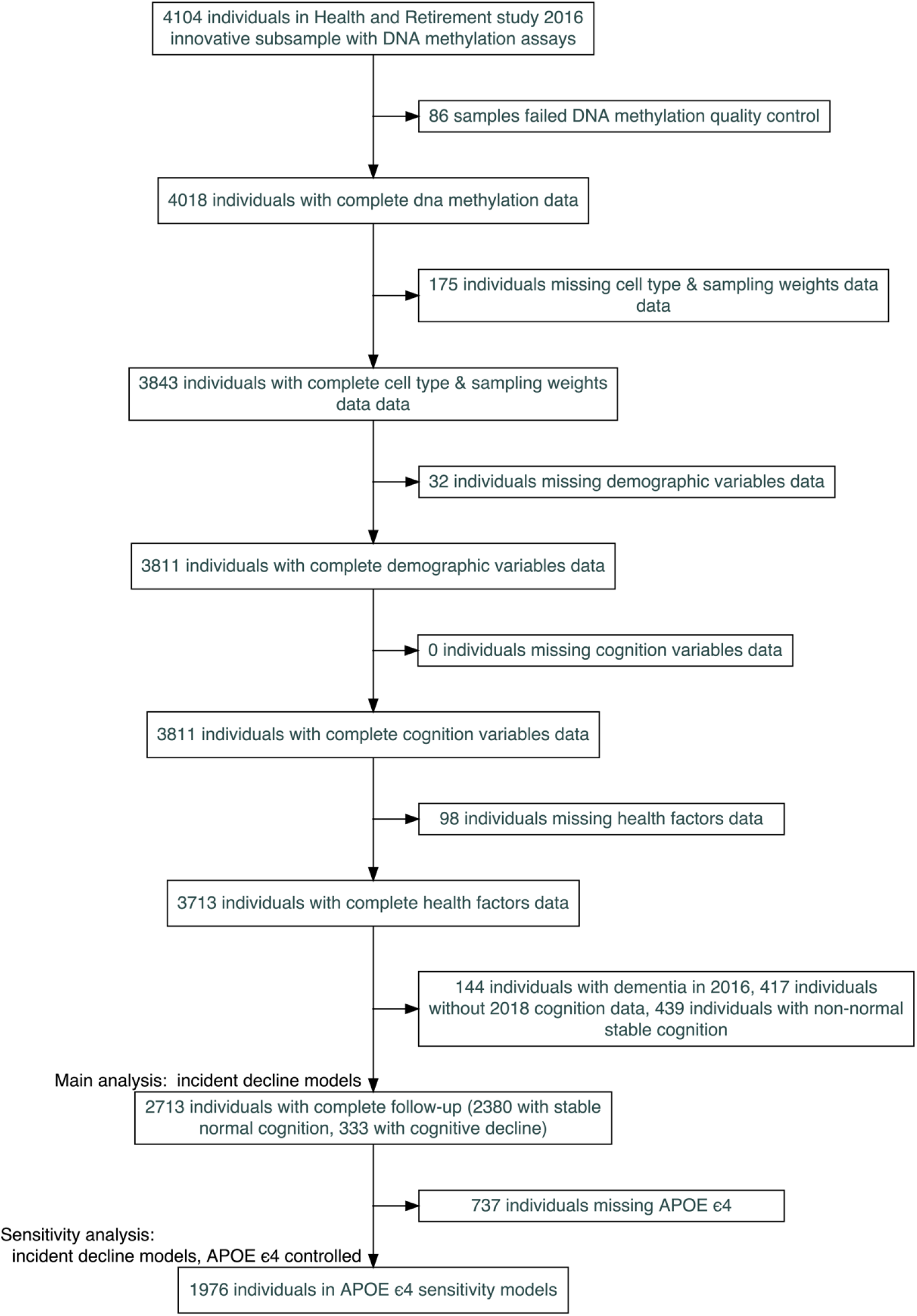
CONSORT (Consolidated Standards of Reporting Trials) diagram for inclusions and exclusions for an analysis of DNA methylation clocks and cognitive decline among a subset of participants in the Health and Retirement Study

Compared to excluded participants, those in the analytic sample were more likely to self-identify as non-Hispanic White, be currently married or partnered, and have more education. They also exhibited fewer chronic conditions, were more likely to exercise at least once per week, and were more often ever-drinkers (**Supplemental Table 1**). Within our final analytic sample, 54% of the participants identified as female. In addition, 86% identified as White, 8.4% as Black, and 7.5% as Hispanic (**Table 1**).

**Table 1.** Survey weighted bivariate descriptive statistics of participant baseline (2016) characteristics in the Health and Retirement Study by cognitive decline status over follow-up (2018).

| Baseline characteristics | Overall<br>(N = 2,713)<sup>1</sup> | Any decline<br>(N = 333)<sup>1</sup> | Normal to normal<br>(N = 2,380)<sup>1</sup> | p-value<sup>2</sup> |
| --- | --- | --- | --- | --- |
| DNA methylation age acceleration (years) |  |  |  |  |
| GrimAge | -0.7 (4.7) | 1.2 (4.5) | -0.8 (4.6) | <0.001 |
| PhenoAge | 0 (7) | 0 (7) | 0 (7) | 0.5 |
| MPOA | -1 (6) | 1 (7) | -1 (6) | <0.001 |
| Horvath | 0.1 (6.0) | -0.4 (6.6) | 0.2 (6.0) | 0.3 |
| Hannum | 0.0 (4.9) | 0.3 (4.9) | 0.0 (4.9) | 0.7 |
| Chronologic age (years) | 68 (8) | 73 (10) | 67 (8) | <0.001 |
| Sex (female) | 54% (0.01) | 48% (0.03) | 54% (0.01) | 0.11 |
| Race |  |  |  | <0.001 |
| White | 86% (0.01) | 77% (0.03) | 87% (0.01) |  |
| Black/African American | 8.4% (0.01) | 17% (0.02) | 7.6% (0.01) |  |
| Other | 5.2% (0.00) | 6.6% (0.02) | 5.0% (0.01) |  |
| Ethnicity (Hispanic) | 7.5% (0.01) | 12% (0.02) | 7.1% (0.01) | 0.008 |
| Educational attainment (years) | 13.77 (2.67) | 12.33 (3.19) | 13.92 (2.57) | <0.001 |
| Marital status (single) | 35% (0.01) | 47% (0.03) | 34% (0.01) | <0.001 |
| Cognitive status |  |  |  | <0.001 |
| Normal cognition | 98% (0.00) | 82% (0.02) | 100% (0.00) |  |
| Cognitively impaired, non-dementia | 1.7% (0.00) | 18% (0.02) | 0% (0.00) |  |
| Exercise status (more than once per week) | 79% (0.01) | 63% (0.03) | 80% (0.01) | <0.001 |
| Drinking status (ever) | 64% (0.01) | 47% (0.03) | 66% (0.01) | <0.001 |
| BMI (kg/m<sup>2</sup>) | 29.0 (6.2) | 28.7 (6.9) | 29.0 (6.1) | 0.4 |
| Chronic conditions (more than one) | 64% (0.01) | 77% (0.03) | 63% (0.01) | <0.001 |
| Smoking status (ever smoker) | 55% (0.01) | 59% (0.03) | 54% (0.01) | 0.15 |
| *APOE* status, any copy of $\epsilon 4$ | 22% (0.01) | 30% (0.03) | 22% (0.01) | 0.009 |
| Granulocytes (%) | 62 (9) | 64 (10) | 62 (9) | <0.001 |
| Lymphocytes (%) | 30 (8) | 28 (9) | 30 (8) | 0.001 |
| Monocytes (%) | 8.54 (2.28) | 8.28 (2.23) | 8.57 (2.28) | 0.042 |
| <sup>1</sup>Mean (SD); frequency% (SE(frequency%)) |  |  |  |  |
| <sup>2</sup>Wilcoxon rank-sum test for complex survey samples; chi-squared test with Rao & Scott's second-order correction |  |  |  |  |

### Bivariate descriptive statistics

Participants who experienced cognitive decline during follow-up were, on average, older and had fewer years of education, compared to those with normal cognition (**Table 1**). They were also more likely to be Black, Hispanic, or single. At baseline, they were less likely to exercise at least once a week or report any current alcohol consumption. Participants who experienced cognitive decline were more likely to having multiple chronic conditions and at least one copy of *APOE* ϵ4 (**Table 1**).

Participants who experienced cognitive decline during follow-up had higher baseline GrimAge (mean = 1.2 years) acceleration compared to those who maintained normal cognitive function (mean = -0.8 years, p < 0.001) (**Table 1 & Supplemental Figure 4**). Participants who experienced cognitive decline during follow-up also had higher baseline MPOA (mean = 1.0 years) acceleration compared to those who maintained normal cognitive function (mean = -1.0 years, p < 0.001). This difference was not observed with other epigenetic clocks. Within the cognitive decline group, those progressing from normal cognition to cognitive impairment non-dementia had lower GrimAge acceleration (mean = 0.94) than those progressing either from normal cognition to dementia (mean = 1.27) or from cognitive impairment non-dementia to dementia (mean = 1.97, **Figure 2**.

**Figure 2.**
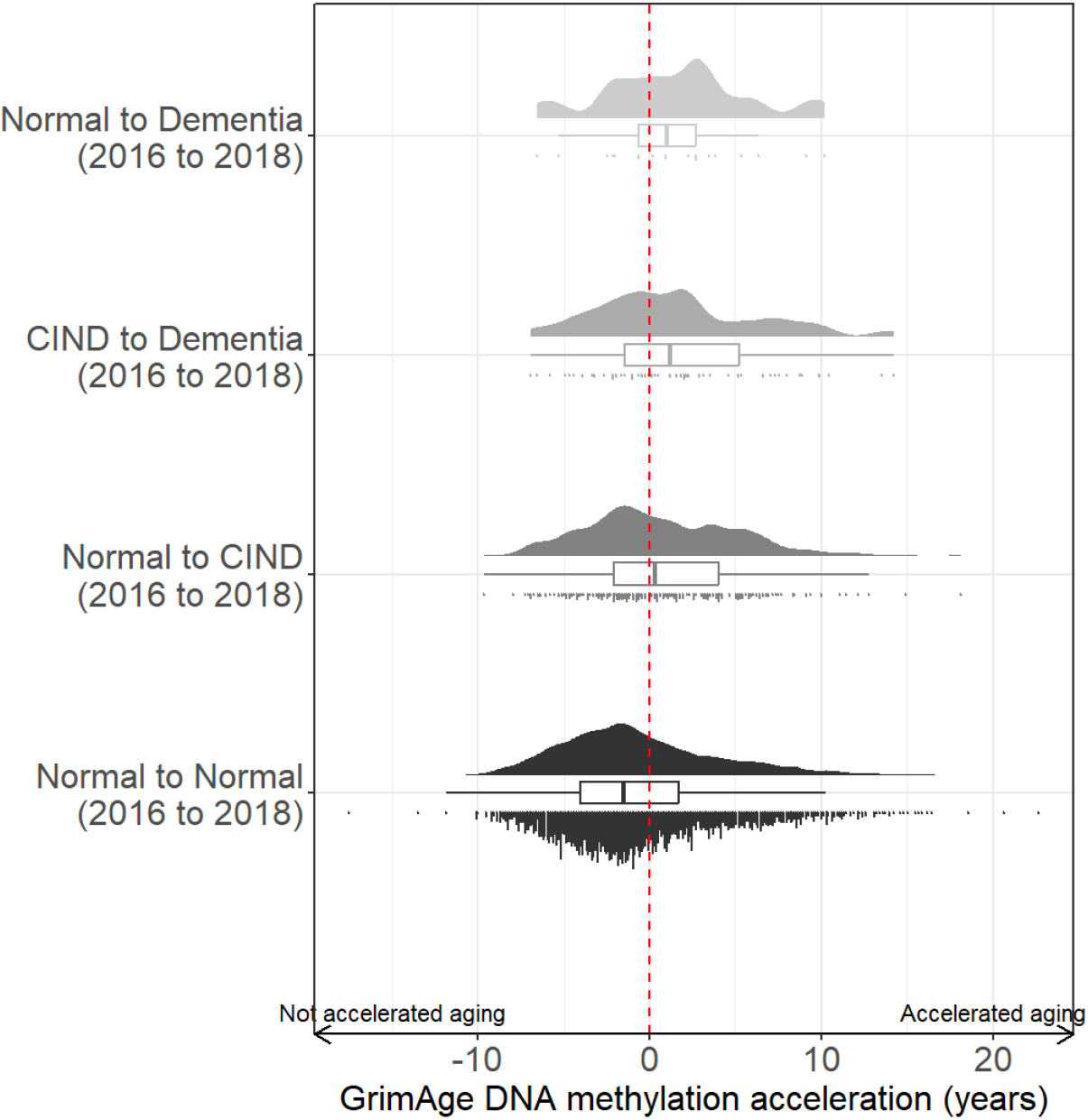
GrimAge acceleration by cognitive decline status (between 2016 and 2018) in the Health and Retirement Study sample with DNA methylation measurements and complete covariate data (N = 2,713). Cognitive impairment non-dementia (CIND).

### Multivariable association between GrimAge acceleration and cognitive decline

Based on *a priori* hypotheses, we chose GrimAge as the primary predictor in our multivariable models. In survey weighted multivariable logistic regressions, we observed that every additional year of increased GrimAge acceleration at baseline was associated with 1.08 (95% CI: 1.05, 1.11, p = 1.0×10^-8^) times higher odds of cognitive decline, after adjusting for cell-type proportions. Controlling for demographic variables slightly attenuated the odds ratio (OR = 1.05, 95% CI: 1.02, 1.09, p = 0.003), and results were similar when we additionally controlling for health variables (OR = 1.05, 95% CI: 1.01, 1.10, p = 0.02). We did not observe a significant interaction between chronological age and GrimAge acceleration in either demographic model or health factors model (**Supplemental Table 2**).

### Sensitivity analyses for parameterization, other clocks, and *APOE* adjustment

When we analyzed the data using a binary parameterization of the exposure, i.e., comparing those with accelerated GrimAge (residuals > 0) against those without (residuals ≤ 0), the results remained consistent (**Table 2**). Full results are available in the supplemental materials. We observed an association between binary MPOA acceleration and odds of cognitive decline (OR = 1.44, 95% CI: 1.03, 2.02, p = 0.03) in the survey weighted fully adjusted (**Supplemental Table 3**). We did not observe associations between the remaining three clocks (PhenoAge, Horvath, Hannum) with cognitive decline.

**Table 2.**
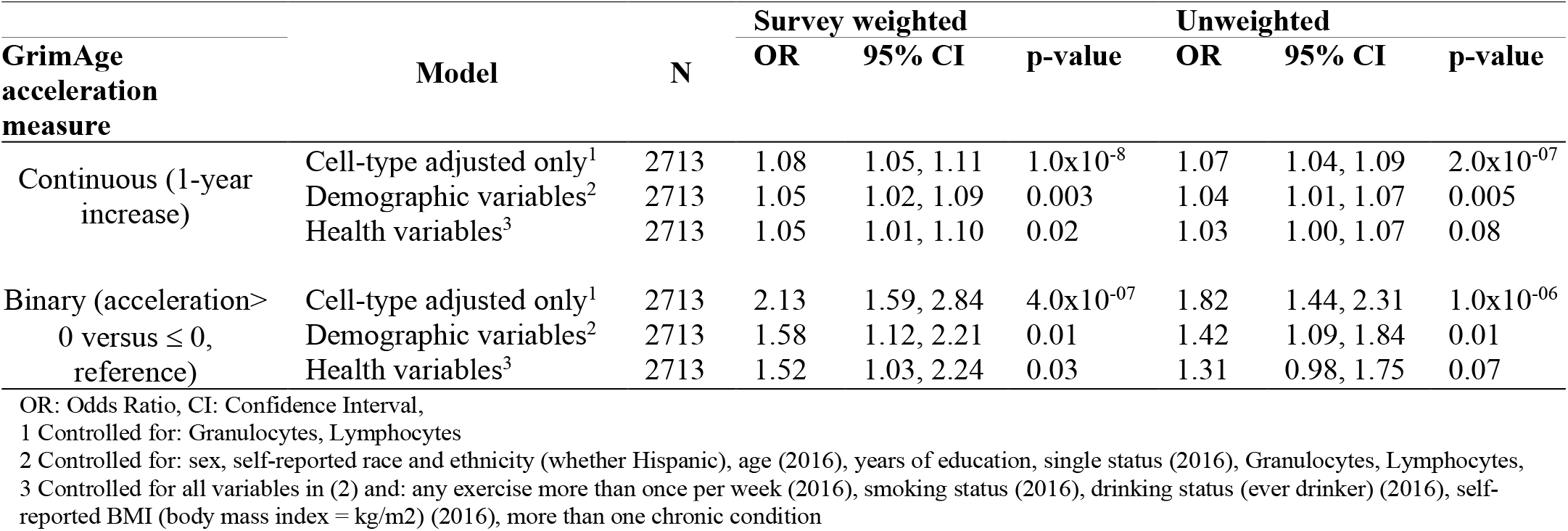
Results from multivariable regression model, predicting any cognitive decline versus stable normal cognition with accelerated epigenetic aging (GrimAge) as the main predictor.

| GrimAge acceleration measure | Model | N | Survey weighted |  |  | Unweighted |  |  |
| --- | --- | --- | --- | --- | --- | --- | --- | --- |
|  |  |  | OR | 95% CI | p-value | OR | 95% CI | p-value |
| Continuous (1-year increase) | Cell-type adjusted only<sup>1</sup> | 2713 | 1.08 | 1.05, 1.11 | $1.0 \times 10^{-8}$ | 1.07 | 1.04, 1.09 | $2.0 \times 10^{-07}$ |
|  | Demographic variables<sup>2</sup> | 2713 | 1.05 | 1.02, 1.09 | 0.003 | 1.04 | 1.01, 1.07 | 0.005 |
|  | Health variables<sup>3</sup> | 2713 | 1.05 | 1.01, 1.10 | 0.02 | 1.03 | 1.00, 1.07 | 0.08 |
| Binary (acceleration > 0 versus ≤ 0, reference) | Cell-type adjusted only<sup>1</sup> | 2713 | 2.13 | 1.59, 2.84 | $4.0 \times 10^{-07}$ | 1.82 | 1.44, 2.31 | $1.0 \times 10^{-06}$ |
|  | Demographic variables<sup>2</sup> | 2713 | 1.58 | 1.12, 2.21 | 0.01 | 1.42 | 1.09, 1.84 | 0.01 |
|  | Health variables<sup>3</sup> | 2713 | 1.52 | 1.03, 2.24 | 0.03 | 1.31 | 0.98, 1.75 | 0.07 |

OR: Odds Ratio, CI: Confidence Interval,
1 Controlled for: Granulocytes, Lymphocytes
2 Controlled for: sex, self-reported race and ethnicity (whether Hispanic), age (2016), years of education, single status (2016), Granulocytes, Lymphocytes,
3 Controlled for all variables in (2) and: any exercise more than once per week (2016), smoking status (2016), drinking status (ever drinker) (2016), self-reported BMI (body mass index = kg/m<sup>2</sup>) (2016), more than one chronic condition

In sensitivity analyses in a subset of participants (N = 1,976) where we were able to control for the *APOE* e4 allele status, results were consistent with the primary analysis that adjusted for other health variables (OR = 1.05, 95% CI: 1.00, 1.10, p = 0.08, **Supplemental Table 4**).

### Sensitivity analyses accounting for loss-to-follow-up

For 417 participants with baseline DNA methylation and cognitive status measures, we were unable to evaluate the cognitive decline due to missing cognitive status at follow-up. Participants missing cognitive data at follow-up had higher baseline GrimAge acceleration (mean: 1.5 years) compared to participants with cognitive data at follow-up (mean: -0.4 years). In addition, participants who were lost to follow-up were more likely to have a smoking history and less likely to have exercise, compared to those who remained in the study (**Supplemental Table 5**).

To address the potential bias introduced by the missing 2018 cognition data, we conducted a sensitivity analysis using inverse probability weighting (**Supplemental Table 6)**. After adjusting for all cell-type, demographic, and health variables, the results mirrored those of the primary multivariable regression (OR = 1.07, 95% CI: 1.04, 1.11, p = 1.0×10^-04^).

### Classification of cognitive status

To assess whether GrimAge acceleration improved classification of cognitive decline, we used receiver operating curve analyses. The AUC for the base model (cell types and demographics) was 0.755. Additionally using GrimAge acceleration resulted in the same AUC (0.755). There was no classification improvement with the inclusion of GrimAge acceleration (DeLong test p-value = 0.3; see **Supplemental Figure 5**).

## Discussion

DNA methylation clocks have emerged as a potential biomarker for cognitive impairment and dementia. However, most longitudinal studies exploring the link between DNA methylation clocks and incident cognitive decline have been limited by small sample sizes (previous N’s: 52 - 578). To address this, we investigated the association between epigenetic clocks and the progression of cognitive decline in a larger sample (N = 2,713) from a longitudinal study focusing on older US adults. We found that participants who experienced cognitive decline over two years of follow up had elevated baseline GrimAge age acceleration, when contrasted with participants maintaining stable cognitive function over follow-up. Specifically, we observed a 1-year increase in baseline GrimAge acceleration was associated with 1.05 (95% CI: (1.01, 1.10)) times higher odds of cognitive decline over follow-up, in fully adjusted models. Moreover, this association remained evident even when accounting for loss-to-follow-up using inverse probability weighting modeling (OR = 1.07, 95% CI: (1.04, 1.11)). In conclusion, our study presents compelling evidence that DNA methylation clocks, particularly GrimAge age acceleration, are biomarkers associated with the progression of cognitive decline in older adults in the United States.

Our study highlights the potential of DNA methylation clocks as biomarkers for impaired cognition. Substantial evidence indicates that alterations in blood DNA methylation patterns are indicative of cognitive dysfunction and senescence of the brain [9,11,29–33]. Notably, a cross-sectional analysis within the Whitehall II imaging sub-study (N = 47) revealed that DNA methylation aging acceleration, as quantified by the Hannum clock, correlated with mean diffusivity and the global fractional anisotropy of the brain [30]. In this study, we noted pronounced differences in cognitive decline exclusively with second-generation phenotypic clocks (GrimAge and MPOA), emphasizing their enhanced ability to predict age-related health declines compared to their first-generation counterparts (Horvath and Hannum). Further endorsement comes from a longitudinal examination within the Irish Longitudinal Study on Aging, where 490 participants monitored over a decade exhibited a marked correlation between GrimAge clock metrics and age-associated cognitive decline [9]. In a similar vein, initial findings from the VITAL-DEP (VITamin D and OmegA-3 TriaL-Depression Endpoint Prevention) study’s pilot cohort (N = 45) observed a significant linkage between GrimAge and rapid declines in overall cognitive function over a two-year span [29]. These studies collectively affirm the viability of DNA methylation clocks, especially GrimAge, as potential biomarkers for early detection of cognitive impairment, which is critical for timely therapeutic intervention.

On the other hand, some prior studies have reported no link between DNA methylation clocks and cognitive decline [12–14,34]. For instance, a case-cohort investigation within the ASPirin in Reducing Events in the Elderly (ASPREE) study, encompassing 160 participants, discerned no differential age acceleration—as measured by Horvath, Hannum, GrimAge, and PhenoAge DNA methylation clocks—between dementia cases and control subjects [13]. Similarly, a cross-sectional analysis utilizing data from 640 participants in the Alzheimer’s Disease Neuroimaging Initiative (ADNI) database also reported no correlations between age acceleration (measured in Horvath, PhenoAge, and GrimAge clocks) and cognitive metrics [34]. Such discrepancies in findings may be attributable to methodological variances, including study design and sample size. Our investigation, leveraging a substantial cohort from the HRS, benefits from a larger sample size, which confers enhanced statistical power.

Our study has several notable strengths that underscore its significance and influence. First, we employed a variety of epigenetic clocks for exposure measurement to examine differences in cognitive decline (**Supplemental Table 3**). This is important as varying associations may exist across these different epigenetic clocks, a notion further supported by our results. Second, compared to earlier longitudinal studies, our sample size is not only larger but also encompasses a nationally representative sample of participants from diverse racial and ethnic groups. This diversity enhances the robustness of our results and conclusions. By incorporating survey-weighting, we ensured that our findings are more representative and thus can be more readily generalized to the broader population of older adults in the US. Moreover, we meticulously accounted for numerous potential confounders in our analysis. We also carried out a sensitivity analysis using inverse probability weighting to tackle potential selection bias.

However, there are certain limitations to our study that must be taken into account when interpreting our results. Notably, the DNA methylation clocks were derived from blood samples rather than brain tissue, which can provide a more direct insight into neural changes and their potential correlation with cognitive decline. Nonetheless, acquiring brain samples from living individuals is not feasible, so this inherent limitation could not be overcome in our study. Many of the epigenetic clocks were designed to be robust across tissues [35]. Additionally, the cognitive status outcome in our study was derived from cognition tests instead of a physician’s diagnosis. This could lead to potential misclassification, and our inability to distinguish between dementia subtypes might result in imprecise conclusions. Furthermore, our study experienced attrition with 417 participants lost to follow-up. This phenomenon could be due to a variety of factors, including withdrawal of participants or mortality stemming from causes unrelated to the study. It is important to consider that such attrition might be non-random and associated with baseline cognitive abilities or rates of epigenetic aging. Lastly, the follow-up period in our analytical sample is limited, spanning only two years of cognitive changes. A more extended follow-up would likely yield clearer and more robust findings.

In conclusion, our study aimed to examine the relationships between DNA methylation age acceleration measured using epigenetic clocks and the onset of cognitive decline. After accounting for specific confounders and potential selection bias, we identified accelerated GrimAge at baseline was associated with cognitive decline over follow-up. By highlighting this association, our study deepens the understanding of the role epigenetic aging clocks might play as reliable predictors for cognitive decline. These insights can pave the way for enhanced prevention, diagnosis, and treatment strategies. By addressing the study’s limitations and leveraging its strengths, the scientific community is better positioned to unravel the intricate ties between epigenetic clocks and cognitive decline.

## Data Availability

All data produced are available online at https://hrsdata.isr.umich.edu/

## Data availability

The Health and Retirement Study data used in this analysis are publicly available (https://hrsdata.isr.umich.edu/). The code used to process the data and produce the analyses are publicly available (https://github.com/bakulskilab).

## Author contributions

FB: Data curation, Formal analysis, Visualization, Writing – original draft; HW: Formal analysis; MF: Writing – original draft; KMB: Conceptualization, Writing – Review & editing, Funding acquisition; EBW: Conceptualization, Funding acquisition; MZ: Writing – Review & editing

## Acknowledgments

We thank the participants and staff of the Health and Retirement Study. The Health and Retirement Study is supported by the National Institute on Aging (U01 AG009740). This analysis was supported by the National Institute on Aging (R01 AG072396, P30 AG072931, R01 AG070897, R01 AG067592).

