## Supplemental materials for "DNA methylation age acceleration is associated with incident cognitive impairment in the Health and Retirement Study"

### Supplementary Materials

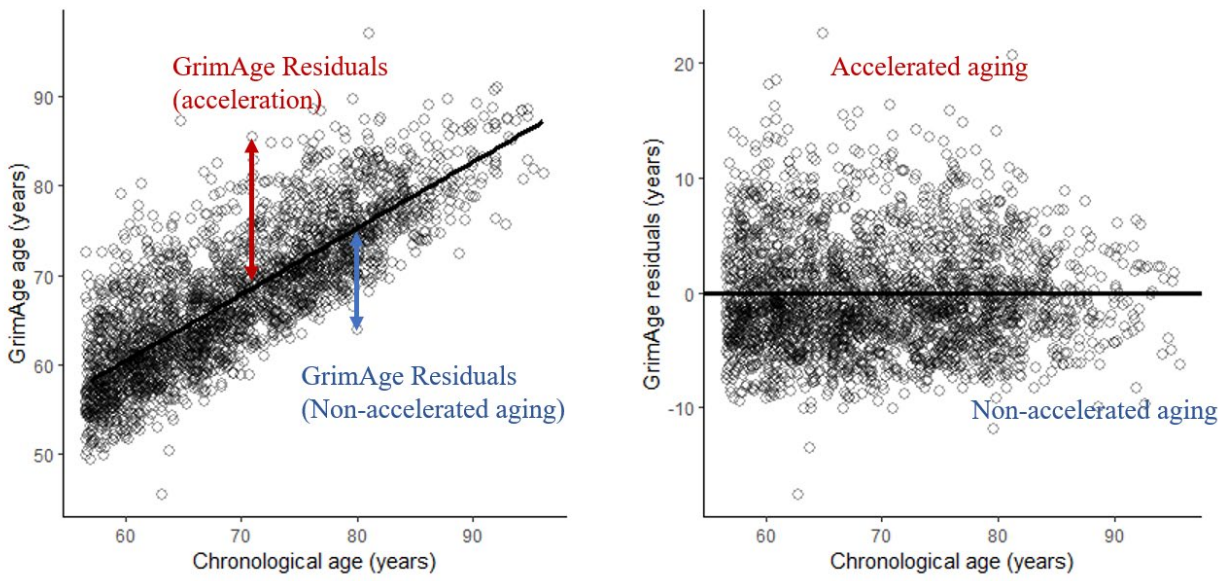


**Supplemental Figure 1.** Pairwise plots of chronological plots with GrimAge and GrimAge residuals (GrimAge acceleration) among the analytic sample with DNA methylation measurements and complete covariate data (N = 2,713)

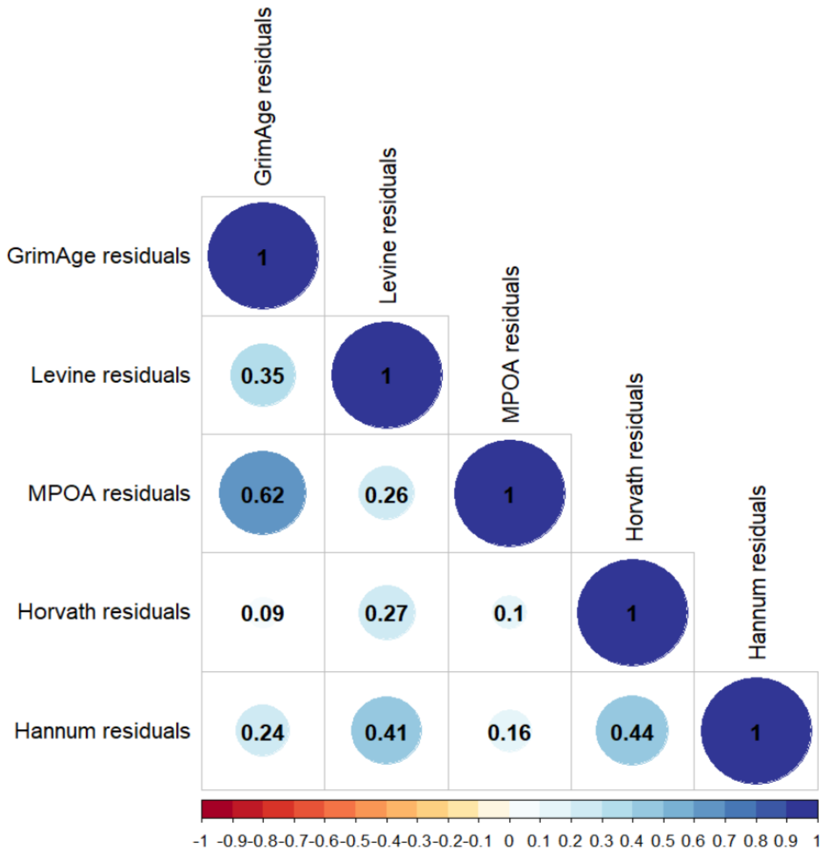


**Supplemental Figure 2.** Pearson correlation plots between epigenetic age acceleration variables. Significant correlations are denoted with colored circle. Correlations are presented in each cell.

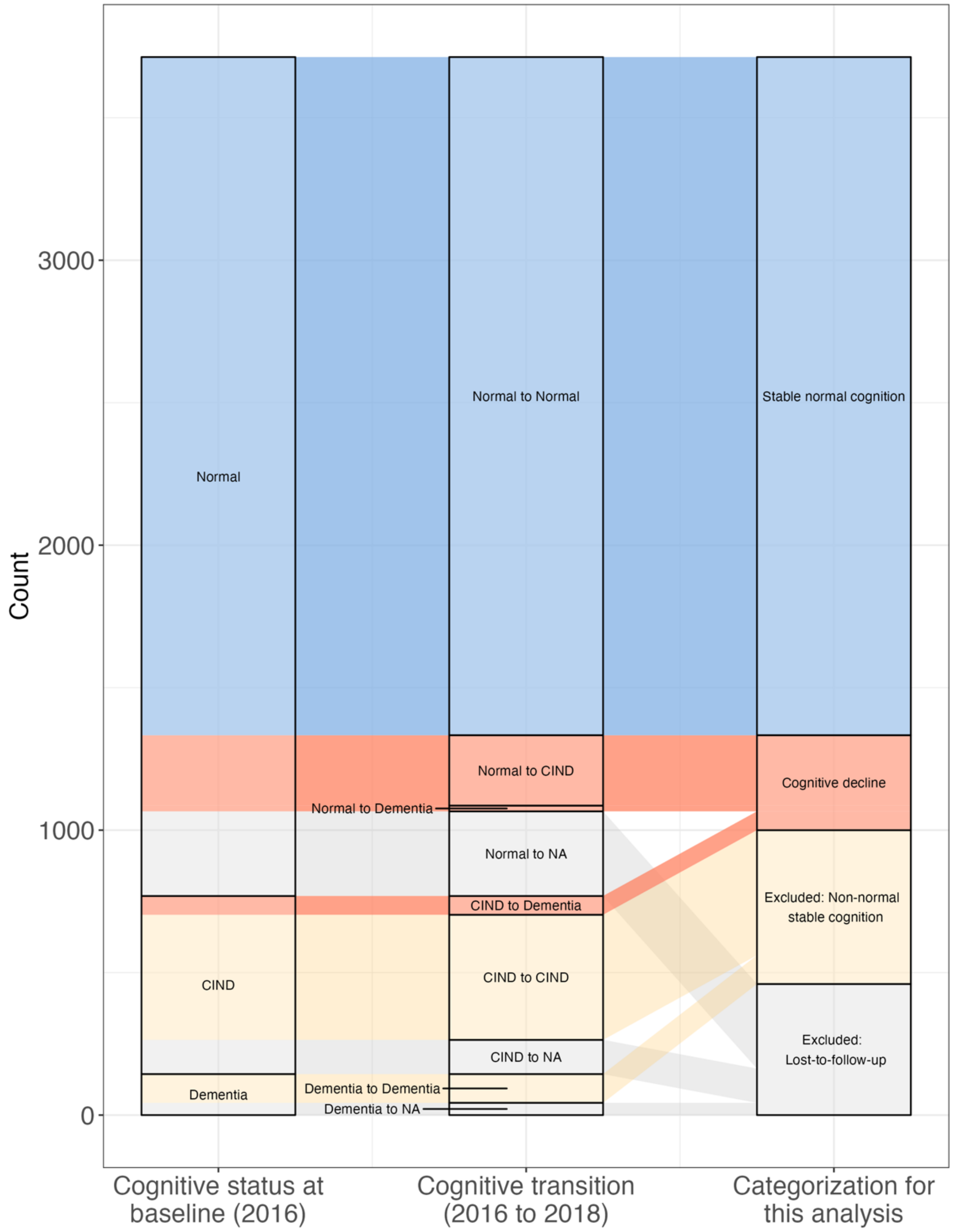


**Supplemental Figure 3.** Construction of the cognitive decline variable for this analysis. Alluvial plot showing cognitive status at baseline (2016) on the left, cognitive transition over follow-up (2018) in the middle, and the cognitive status operationalization for this analysis on the right. Blue shows participants operationalized with stable normal cognition. Orange shows participants operationalized with cognitive decline. Yellow shows participants excluded due to non-declining, non-normal cognition states. Grey shows participants excluded due to loss-to-follow-up.

*Note.* CIND: Cognitive impairment non-dementia, NA: missing

**Supplemental Table 1**. Among the overall sample of Health and Retirement Study participants with DNA methylation data, we compare the included analytic sample to the excluded sample

| **Baseline characteristics** | **Overall N = 4,018**[1] | **Excluded N = 1,305**[1] | **Included N = 2,713**[1] | **p-value**[2] |
|---|---|---|---|---|
| Sex (Female) | 2,349 (58%) | 757 (58%) | 1,592 (59%) | 0.7 |
| Race | | | | <0.001 |
| White | 3,013 (75%) | 881 (68%) | 2,132 (79%) | |
| Black/African American | 674 (17%) | 272 (21%) | 402 (15%) | |
| Other | 318 (7.9%) | 139 (11%) | 179 (6.6%) | |
| Unknown | 13 | 13 | 0 | |
| Ethnicity | | | | <0.001 |
| Not Hispanic | 3,448 (86%) | 1,037 (80%) | 2,411 (89%) | |
| Hispanic | 567 (14%) | 265 (20%) | 302 (11%) | |
| Unknown | 3 | 3 | 0 | |
| Chronologic age (years) | 70 (10) | 70 (11) | 70 (9) | 0.054 |
| Educational attainment (years) | 12.8 (3.2) | 11.6 (3.6) | 13.4 (2.8) | <0.001 |
| Unknown | 19 | 19 | 0 | |
| Marital status | | | | <0.001 |
| Married or partnered | 2,484 (62%) | 736 (57%) | 1,748 (64%) | |
| Single | 1,531 (38%) | 566 (43%) | 965 (36%) | |
| Unknown | 3 | 3 | 0 | |
| Cognitive status | | | | <0.001 |
| Normal | 3,183 (79%) | 536 (41%) | 2,647 (98%) | |
| Cognitive impairment non-dementia | 683 (17%) | 617 (47%) | 66 (2.4%) | |
| Dementia | 152 (3.8%) | 152 (12%) | 0 (0%) | |
| Exercise status (more than once per week) | 2,814 (71%) | 782 (62%) | 2,032 (75%) | <0.001 |
| Unknown | 39 | 39 | 0 | |
| Drinking status (ever) | 2,234 (56%) | 612 (47%) | 1,622 (60%) | <0.001 |
| Unknown | 1 | 1 | 0 | |
| BMI ($kg/m^2$) | 28.9 (6.3) | 28.6 (6.4) | 29.0 (6.2) | 0.046 |
| Unknown | 43 | 43 | 0 | |
| Chronic conditions (more than one) | 2,848 (71%) | 968 (74%) | 1,880 (69%) | 0.001 |
| Smoking status | | | | 0.3 |
| Never smoker | 1,764 (44%) | 550 (43%) | 1,214 (45%) | |
| Ever smoker | 2,230 (56%) | 731 (57%) | 1,499 (55%) | |
| Unknown | 24 | 24 | 0 | |
| *APOE* status, any copy of e4 | 698 (24%) | 230 (26%) | 468 (24%) | 0.2 |
| Unknown | 1,157 | 420 | 737 | |
| Granulocytes (%) | 62 (10) | 62 (10) | 62 (9) | 0.9 |
| Unknown | 33 | 33 | 0 | |
| Lymphocytes (%) | 30 (9) | 30 (10) | 30 (9) | 0.7 |
| Unknown | 33 | 33 | 0 | |
| Monocytes (%) | 8.52 (2.43) | 8.49 (2.65) | 8.53 (2.32) | 0.6 |
| Unknown | 33 | 33 | 0 | |

[1]n (%); Mean (SD)

[2]Pearson's Chi-squared test; Welch Two Sample t-test

**Supplemental Table 2.** Results from multivariable regression model, predicting any cognitive decline versus stable normal cognition with continuous DNA methylation age acceleration (GrimAge) as the main predictor and chronological age as effect modifier

| | **Survey weighted OR (95% CI)** | | **Unweighted OR (95% CI)** | |
|---|---|---|---|---|
| | Demographic model[1] | Health variables model[2] | Demographic model[1] | Health variables model[2] |
| Continuous GrimAge acceleration | 1.29 (1.00, 1.65) | 1.32 (1.02, 1.70) | 1.20 (0.97, 1.48) | 1.21 (0.97, 1.52) |
| Chronological age | 1.09 (1.06, 1.11) | 1.08 (1.06, 1.10) | 1.09 (1.07, 1.10) | 1.08 (1.06, 1.10) |
| Continuous GrimAge acceleration * Chronological age | 1.00 (0.99, 1.00) | 1.00 (0.99, 1.00) | 1.00 (1.00, 1.00) | 1.00 (0.99, 1.00) |

OR: Odds Ratio, CI: Confidence Interval,
1 Controlled for: sex, self-reported race and ethnicity (whether Hispanic), age (2016), years of education, single status (2016), Granulocytes, Lymphocytes,
2 Controlled for all variables in (2) and: any exercise more than once per week (2016), smoking status (2016), drinking status (ever drinker) (2016), self-reported BMI (body mass index = kg/m2) (2016), more than one chronic condition

**Supplemental Table 3.** Results from multivariable regression model, predicting any cognitive decline versus stable normal cognition with other DNA methylation age acceleration as predictors

| DNA methylation acceleration measures | Model | N | Survey weighted | | | Unweighted | | |
|---|---|---|---|---|---|---|---|---|
| | | | OR | 95% CI | p-value | OR | 95% CI | p-value |
| **MPOA** | | | | | | | | |
| Continuous (1-year increase) | Cell-type adjusted only[1] | 2713 | 1.04 | 1.01, 1.07 | 0.002 | 1.03 | 1.01, 1.05 | 0.01 |
| | Demographic variables[2] | 2713 | 1.02 | 0.99, 1.04 | 0.21 | 1.01 | 0.99, 1.03 | 0.36 |
| | Health variables[3] | 2713 | 1.01 | 0.98, 1.04 | 0.53 | 1.00 | 0.99, 1.02 | 1.00 |
| Binary (acceleration> 0 versus ≤ 0, reference) | Cell-type adjusted only[1] | 2713 | 1.91 | 1.40, 2.61 | $4.0x10^{-5}$ | 1.52 | 1.20, 1.94 | 0.001 |
| | Demographic variables[2] | 2713 | 1.52 | 1.11, 2.09 | 0.01 | 1.27 | 0.98, 1.65 | 0.07 |
| | Health variables[3] | 2713 | 1.44 | 1.03, 2.02 | 0.03 | 1.18 | 0.89, 1.56 | 0.25 |
| **PhenoAge** | | | | | | | | |
| Continuous (1-year increase) | Cell-type adjusted only[1] | 2713 | 1.00 | 0.98, 1.02 | 0.92 | 1.00 | 0.99, 1.02 | 0.64 |
| | Demographic variables[2] | 2713 | 1.00 | 0.98, 1.02 | 0.96 | 1.00 | 0.99, 1.02 | 0.80 |
| | Health variables[3] | 2713 | 1.00 | 0.97, 1.02 | 0.69 | 1.00 | 0.98 1.02 | 0.92 |
| Binary (acceleration> 0 versus ≤ 0, reference) | Cell-type adjusted only[1] | 2713 | 0.93 | 0.69, 1.26 | 0.65 | 1.07 | 0.85, 1.36 | 0.56 |
| | Demographic variables[2] | 2713 | 0.93 | 0.68, 1.27 | 0.63 | 1.04 | 0.81, 1.33 | 0.78 |
| | Health variables[3] | 2713 | 0.89 | 0.65, 1.22 | 0.47 | 1.01 | 0.78, 1.30 | 0.94 |
| **Horvath** | | | | | | | | |
| Continuous (1-year increase) | Cell-type adjusted only[1] | 2713 | 0.98 | 0.96, 1.01 | 0.15 | 0.99 | 0.97, 1.00 | 0.13 |
| | Demographic variables[2] | 2713 | 0.98 | 0.96, 1.01 | 0.14 | 0.98 | 0.97, 1.00 | 0.10 |
| | Health variables[3] | 2713 | 0.98 | 0.96, 1.00 | 0.12 | 0.98 | 0.97, 1.00 | 0.09 |
| Binary (acceleration> 0 versus ≤ 0, reference) | Cell-type adjusted only[1] | 2713 | 0.84 | 0.63, 1.12 | 0.24 | 0.86 | 0.67, 1.09 | 0.21 |
| | Demographic variables[2] | 2713 | 0.82 | 0.61, 1.11 | 0.20 | 0.86 | 0.67, 1.10 | 0.22 |
| | Health variables[3] | 2713 | 0.81 | 0.60, 1.10 | 0.17 | 0.85 | 0.66, 1.08 | 0.19 |
| **Hannum** | | | | | | | | |
| Continuous (1-year increase) | Cell-type adjusted only[1] | 2713 | 1.00 | 0.97, 1.03 | 1.00 | 0.99 | 0.96, 1.01 | 2e-07 |
| | Demographic variables[2] | 2713 | 1.01 | 0.97, 1.03 | 0.66 | 0.99 | 0.97, 1.02 | 0.005 |
| | Health variables[3] | 2713 | 1.00 | 0.97, 1.03 | 0.92 | 0.99 | 0.96, 1.01 | 0.08 |

| | | | | | | | | |
|---|---|---|---|---|---|---|---|---|
| Binary (acceleration> 0 versus ≤ 0, reference) | Cell-type adjusted only[1] | 2713 | 0.77 | 0.57, 1.05 | 0.10 | 0.72 | 0.56, 0.91 | 0.01 |
| | Demographic variables[2] | 2713 | 0.81 | 0.59, 1.10 | 0.17 | 0.72 | 0.56, 0.93 | 0.01 |
| | Health variables[3] | 2713 | 0.76 | 0.56, 1.04 | 0.09 | 0.67 | 0.52, 0.87 | 0.003 |

OR: Odds Ratio, CI: Confidence Interval,

1 Controlled for: Granulocytes, Lymphocytes

2 Controlled for: sex, self-reported race and ethnicity (whether Hispanic), age (2016), years of education, single status (2016), Granulocytes, Lymphocytes,

3 Controlled for all variables in (2) and: any exercise more than once per week (2016), smoking status (2016), drinking status (ever drinker) (2016), self-reported BMI (body mass index = kg/m2) (2016), more than one chronic condition

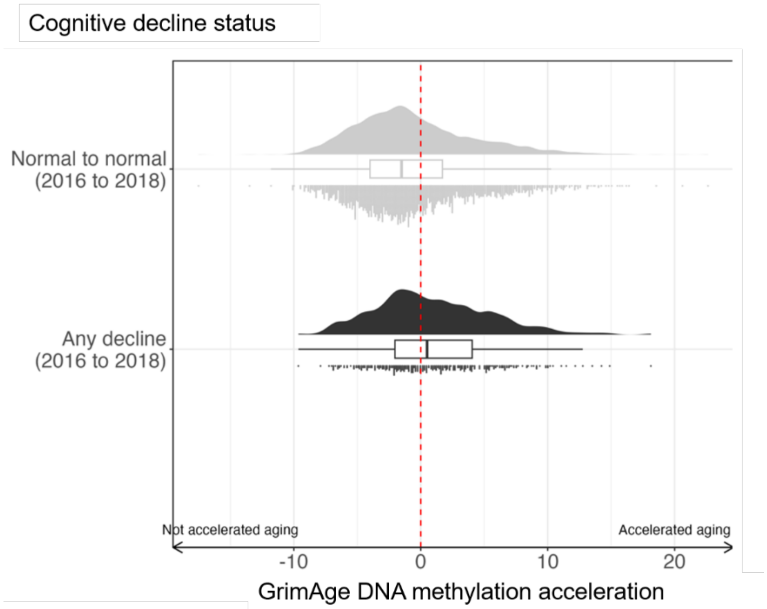


**Supplemental Figure 4.** GrimAge acceleration by cognitive decline status (between 2016 and 2018) in the Health and Retirement Study sample with DNA methylation measurements and complete covariate data (N = 2,713).

**Supplemental Table 4.** Results from sensitivity analyses also controlling for *APOE* e4 status (any versus no alleles of e4)

| GrimAge acceleration measure | Model | N | Survey weighted | | | Unweighted | | |
|---|---|---|---|---|---|---|---|---|
| | | | OR | 95% CI | P value | OR | 95% CI | P value |
| Continuous (1 year) | *APOE* sensitivity models | 1976 | 1.05 | 1.00, 1.10 | 0.08 | 1.03 | 0.99, 1.07 | 0.17 |
| Binary (acceleration> 0 versus reference ≤ 0) | *APOE* sensitivity models | 1976 | 1.28 | 0.85, 1.94 | 0.24 | 1.24 | 0.88, 1.74 | 0.21 |

OR: Odds Ratio, CI: Confidence Interval,

1 Controlled for: sex, self-reported race and ethnicity (whether Hispanic), age (2016), years of education, single status (2016), Granulocytes, Lymphocytes, any exercise more than once per week (2016), smoking status (2016), drinking status (ever-drinker) (2016), self-reported BMI (body mass index = kg/m2) (2016), more than one chronic condition, and *APOE* e4

**Supplemental Table 5.** Analytic subsample versus subset of the Health and Retirement study with epigenetic data and with complete covariate data in 2016

| **Baseline characteristics** | **Overall N = 3,713**[1] | **Excluded due to missing 2018 cognitive status, N = 417**[1] | **Included in analytic sample (Cognitive decline or stable normal cognition), N = 2,713**[1] | **Non-normal stable cognition, N = 583**[1] | **p-value**[2] |
|---|---|---|---|---|---|
| DNA methylation age acceleration | | | | | |
| GrimAge | 0.0 (4.8) | 1.5 (4.9) | -0.4 (4.7) | 0.7 (4.8) | <0.001 |
| MPOA | 0 (6) | 1 (7) | 0 (6) | 1 (7) | <0.001 |
| PhenoAge | 0 (7) | 1 (7) | 0 (7) | 0 (7) | <0.001 |
| Horvath | 0.0 (6.4) | 0.3 (6.8) | 0.0 (6.3) | -0.4 (6.7) | 0.007 |
| Hannum | 0.0 (5.2) | 1.0 (5.8) | -0.1 (5.1) | -0.3 (5.3) | <0.001 |
| Chronologic age (years) | 70 (9) | 71 (11) | 70 (9) | 74 (10) | <0.001 |
| Sex (Female) | 2,139 (58%) | 222 (53%) | 1,592 (59%) | 325 (56%) | 0.068 |
| Race | | | | | <0.001 |
| White | 2,808 (76%) | 317 (76%) | 2,132 (79%) | 359 (62%) | |
| Black/African American | 626 (17%) | 67 (16%) | 402 (15%) | 157 (27%) | |
| Other | 279 (7.5%) | 33 (7.9%) | 179 (6.6%) | 67 (11%) | |
| Ethnicity (Hispanic) | 482 (13%) | 52 (12%) | 302 (11%) | 128 (22%) | <0.001 |
| Educational attainment (years) | 12.9 (3.2) | 12.3 (2.9) | 13.4 (2.8) | 10.7 (3.8) | <0.001 |
| Marital status (single) | 1,442 (39%) | 189 (45%) | 965 (36%) | 288 (49%) | <0.001 |
| Cognitive status | | | | | <0.001 |
| Normal | 2,944 (79%) | 297 (71%) | 2,647 (98%) | 0 (0%) | |
| Cognitive impairment non-dementia | 625 (17%) | 120 (29%) | 66 (2.4%) | 439 (75%) | |
| Dementia | 144 (3.9%) | 0 (0%) | 0 (0%) | 144 (25%) | |
| Exercise status (more than once per week) | 2,616 (70%) | 232 (56%) | 2,032 (75%) | 352 (60%) | <0.001 |
| Drinking status (ever) | 2,053 (55%) | 200 (48%) | 1,622 (60%) | 231 (40%) | <0.001 |
| BMI ($kg/m^2$) | 28.9 (6.2) | 28.2 (6.5) | 29.0 (6.2) | 28.4 (6.0) | 0.002 |
| Chronic conditions (more than one) | 2,652 (71%) | 316 (76%) | 1,880 (69%) | 456 (78%) | <0.001 |
| Smoking status (ever smoker) | 2,091 (56%) | 266 (64%) | 1,499 (55%) | 326 (56%) | 0.005 |
| *APOE* status, any copy of e4 | 654 (24%) | 62 (24%) | 468 (24%) | 124 (28%) | 0.2 |
| Unknown | 1,029 | 155 | 737 | 137 | |
| Granulocytes (%) | 62 (10) | 63 (11) | 62 (9) | 61 (10) | 0.031 |
| Lymphocytes (%) | 30 (9) | 29 (11) | 30 (9) | 30 (10) | 0.004 |
| Monocytes (%) | 8.56 (2.43) | 8.75 (3.01) | 8.53 (2.32) | 8.53 (2.45) | 0.8 |

[1]Mean (SD); n (%)

[2]Kruskal-Wallis rank sum test; Pearson's Chi-squared test

**Supplemental Table 6.** Results from inverse probability weighted analysis with both exposure (accelerated GrimAge) and censoring (no cognitive status in 2018) weights (multivariable logistic models with dependent variable any cognitive decline)

| GrimAge acceleration measure | Model | N | Survey*IPW weighted | | | IPW weighted only | | |
|---|---|---|---|---|---|---|---|---|
| | | | **OR** | **95% CI** | **P value** | **OR** | **95% CI** | **P value** |
| Continuous (1 year) | Demographic variables | 2713 | 1.05 | 1.02, 1.08 | 0.002 | 1.02 | 0.98, 1.05 | 0.3 |
| | Health variables (never vs ever smoker) | 2713 | 1.07 | 1.04, 1.11 | 1e-04 | 1.04 | 1.01, 1.08 | 0.02 |
| Binary (acceleration > 0 versus reference≤ 0) | Demographic variables | 2713 | 1.47 | 1.06, 2.04 | 0.02 | 1.29 | 0.99, 1.68 | 0.06 |
| | Health variables (never vs ever smoker) | 2713 | 1.36 | 0.95, 1.93 | 0.09 | 1.25 | 0.94, 1.68 | 0.12 |

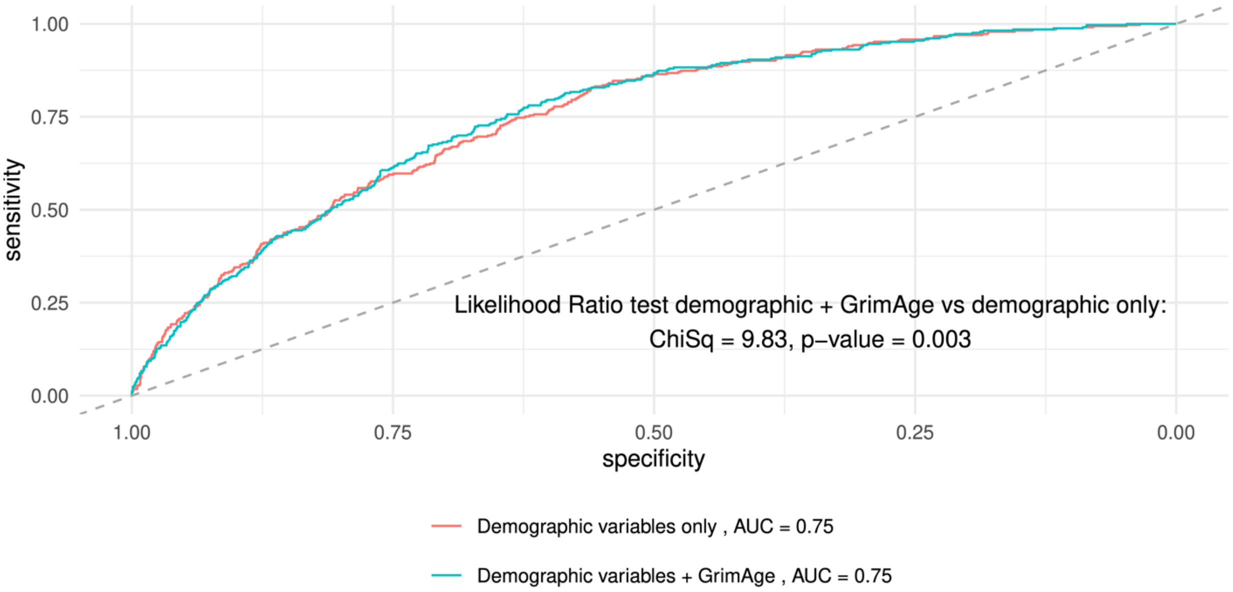


**Supplemental Figure 5.** Receiver operating curve plot from multivariable logistic regressions predicting any cognitive decline when using cell types and demographic variables only (red), or GrimAge residuals + cell types and demographic variables (blue)
